# Optimized navigator-based correction of breathing-induced B_0_ field fluctuations in multi-echo gradient-echo imaging of the spinal cord

**DOI:** 10.1101/2024.12.05.24318389

**Authors:** Laura Beghini, Silvan Büeler, Martina D. Liechti, Alexander Jaffray, Gergely David, S. Johanna Vannesjo

## Abstract

**Purpose:** Multi-echo gradient-echo (ME-GRE) imaging in the spinal cord is susceptible to breathing-induced B_0_ field fluctuations due to the proximity of the lungs, leading to ghosting artifacts. A navigator readout can be used to monitor the fluctuations; however, standard navigator processing often fails in the spinal cord. Here, we introduce navigator processing tailored specifically for spinal cord imaging.

**Methods:** ME-GRE data covering all spinal cord regions were acquired in six healthy volunteers during free breathing at 3T. Centerline navigator readouts and respiratory belt recordings were collected during the acquisitions. The navigator processing included a Fast Fourier Transform and subsequent interval selection targeting the spinal cord, as well as SNR-weighted averaging over samples and coils on the complex data. Furthermore, a phase unwrapping algorithm making use of the belt recordings was developed. Imaging data were corrected by phase demodulation before image reconstruction.

**Results:** B_0_ field fluctuations and ghosting artifacts were largest in the lower cervical and upper lumbosacral cord (∼5 Hz std), close to the edges of the lungs. Image reconstruction based on optimized navigator correction improved visual image quality and quantitative metrics (SNR, CNR, ghosting) in all regions of the spinal cord. The improvement was largest in regions with large field fluctuations (group-averaged increase in SNR/CNR of up to 29%/37% in single-echo images).

**Conclusions:** Optimized navigator-based correction can reduce ghosting artifacts and increase SNR/CNR in anatomical ME-GRE of the spinal cord. The enhanced image quality and ease of implementation across sites makes the technique attractive for clinical and scientific applications.

## Introduction

Multi-echo gradient-echo (ME-GRE) sequences are commonly used in spinal cord imaging, as they provide good contrast between grey matter (GM), white matter (WM), and cerebrospinal fluid (CSF)^1,2^. In addition, they find applications in quantitative imaging, such as quantitative susceptibility mapping (QSM), T2* relaxometry, and myelin water imaging^3^. However, these sequences are sensitive to B_0_ field variations caused by magnetic susceptibility differences between tissue and air^4,5^. Due to the changing air volume in the lungs during breathing, the B_0_ field fluctuates over time^6^, leading to ghosting artifacts and signal loss. Various approaches have been developed to address breathing-induced B_0_ field fluctuations^7–18^. However, most of these studies have targeted brain imaging ^7–14^. Although breathing-induced B_0_ field fluctuations can be observed in the brain^9^, they are more pronounced in the spinal cord^15^ due to its proximity to the lungs. A few studies have explored corrections specifically for spinal cord imaging^16–18^, however, in practice, such corrections are rarely applied, despite the potential gain in image quality.

One key challenge is achieving accurate detection of the field fluctuations within the spinal cord. Previous correction methods relied on recordings from a respiratory belt to track the breathing cycle^16–18^, combined with subject- and session-specific calibration data to correlate the respiratory trace with the B_0_ field fluctuations. However, the correlation may suffer from limited accuracy, especially if the breathing pattern varies during the acquisition. In brain imaging, the most common approach is to use navigator readouts^9,10,14,19,20^, which can directly measure field fluctuations within the tissue and do not require additional hardware. However, their application to the spinal cord is complicated by the lower signal-to-noise ratio (SNR), larger SNR variations between receiver coils, and higher spatial variability in breathing-induced fields. Furthermore, large field fluctuations can cause phase wrapping in the navigator phase estimates, leading to erroneous field estimates. Under these circumstances, standard navigator implementations often fail in the spinal cord and may even exacerbate the artifacts.

In this study, we propose a navigator processing pipeline specifically tailored to address breathing-induced B_0_ field fluctuations in the spinal cord. The pipeline takes data from a standard single-line navigator readout, with the option to combine this data with recordings from a respiratory belt to correct for phase wrapping. The correction is applied retrospectively as a phase demodulation on the acquired k-space data. We assess the impact of the correction in ME-GRE imaging of the cervical, thoracic, and lumbosacral spinal cord at 3T. In addition, we measure the amplitude of the field fluctuations along the spine, extending previously published characterizations^15,21^ to the lumbosacral cord. Preliminary results of this work have previously been published in conference abstracts^22,23^.

## Methods

Six healthy volunteers (2 females, 4 males, age (mean ± std, range): 37.9±12.0y, 24-55y) participated in this study. The study was approved by the local ethics committee (Kantonale Ethikkommission Zürich, BASEC ID: 2019-00074) and conducted in accordance with the Declaration of Helsinki. Written informed consent was obtained from all participants.

### Data acquisition

Scanning was performed on a 3T Siemens Prisma MR system (Siemens Healthineers, Erlangen, Germany) using the body transmit coil and two receive coils: the standard 32-channel spine coil and the 64-channel head and neck coil. Both receive coils were used for imaging the cervical and thoracic cord, while only the spine coil was used for the lumbosacral cord. Volunteers were positioned and instructed as described by Büeler et al.^24^ to minimize motion artifacts. All sequences were acquired during free breathing. Three localizer scans were acquired for the cervical, thoracic, and lumbosacral spinal cord, respectively. An additional sagittal T2-weighted turbo spin echo sequence was acquired as an anatomical reference of the lumbosacral cord^24^.

Three 2D ME-GRE acquisitions were obtained in a counterbalanced order across subjects, covering the cervical, thoracic, and lumbosacral spinal cord, respectively (Figure 1). Seven slabs, each consisting of two axial-oblique slices with no gap, were acquired in both the cervical and thoracic spinal cord and were individually centered at mid-vertebral levels (C2-T1 and T2-T8, respectively) to reduce artifacts due to static B_0_ field inhomogeneities. Each slab was angulated perpendicularly to the spinal cord to reduce partial volume effects. The misalignment between vertebral and spinal levels in the lumbosacral cord ^25^ precluded the use of vertebral levels as neuroanatomical landmarks. Moreover, the spinal curvature is lower in this region. Therefore,18 axial-oblique slices with no gap were acquired within a single slab, which was positioned based on the sagittal T2-weighted image to cover the entire lumbosacral cord as previously described^24,26^.

**Figure 1:**
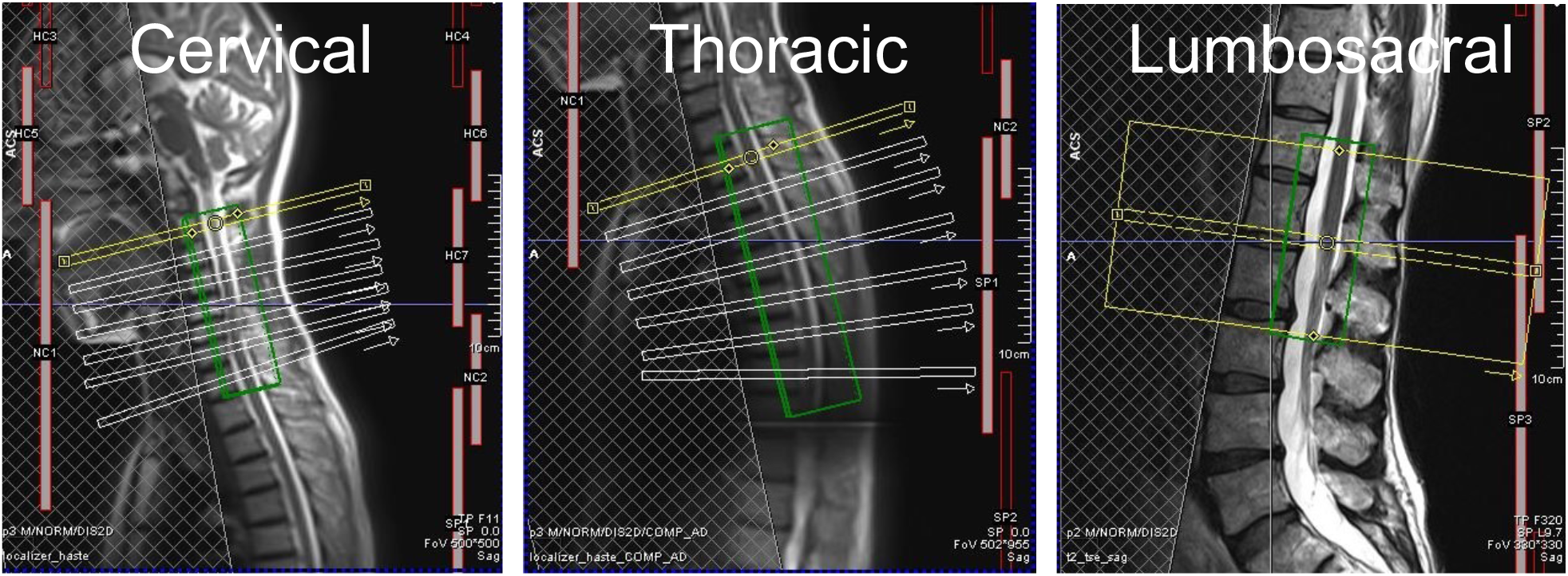
Slice positioning of the 2D ME-GRE sequence in the cervical, thoracic, and lumbosacral spinal cord. The shimming box, outlined in green, encompassed a volume that extended slightly across the spinal canal in the left-right and anterior-posterior direction. A saturation band was placed anteriorly to the vertebral column to suppress signal from the head and neck region, thorax, and abdomen.

The ME-GRE sequences were acquired with the following parameters: 5 mm slice thickness, 0.5×0.5 mm^2^ in-plane resolution, 192×192 mm^2^ in-plane FOV, 4 echoes, TE 6.86/10.86/14.86/18.86 ms (4 ms echo spacing), TR 700 ms (cervical, thoracic) and 899 ms (lumbosacral), flip angle 38° (cervical, thoracic) and 44° (lumbosacral), 2x acceleration factor, 3 repetitions, anterior-posterior (AP) phase encoding, acquisition time 7:12 min (cervical, thoracic) and 9:14 min (lumbosacral). In two subjects, an additional ME-GRE sequence with a longer TR (2 s) was acquired in the lumbosacral cord to investigate the TR-dependence of the navigator correction. After the last echo in each TR for each slice, a navigator was acquired by reading out a single line along the readout (left-right) direction through the center of k-space. This was achieved by using the “phase stabilization” option in the standard GRE sequence provided by the vendor (Syngo v. VE11C), which required minor sequence modification to activate the option. The sequence also included the readout of a noise profile without signal excitation. A trace from the vendor-provided respiratory belt was recorded for all subjects during the ME-GRE acquisitions. A fully sampled, low-resolution GRE reference scan was also acquired using the same slice geometry, with the following parameters: 2×2 mm^2^ in-plane resolution, 256×208 mm^2^ in-plane FOV, TE 3.06 ms, TR 600 ms, flip angle 25°, single repetition, AP phase encoding, and 1:04 min acquisition time.

### Navigator processing

Navigator processing, data correction and image reconstruction were performed offline. The respiratory belt recordings and the raw MR data were synchronized in Matlab (The MathWorks, Natick, Massachusetts, USA) using the PhysIO toolbox^27^. The MRI data were then converted to the ISMRMRD format^28^ and imported in Julia^29^ (v1.8.5) using the MRIReco^30^ (v0.7.1) package. An additional Julia package called MRINavigator (v0.1.1, https://github.com/NordicMRspine/MRINavigator.jl) was developed for this study, including all functions needed for navigator data extraction, navigator processing, and raw data correction along with documentation.

An overview of the navigator processing pipelines is provided in Figure 2. One of the main challenges with phase navigators in spinal cord imaging is the risk of corrupted phase estimates due to low SNR. The navigator processing was therefore designed to yield robust phase estimates by combining data from different sample points, read-out lines, and coils. The simplest correction approach assumed spatially homogeneous field fluctuations within each slice, allowing for estimating field fluctuations from the data at the center of k-space (*k nav*). However, this assumption is not a good approximation of the actual field fluctuations in the spinal cord^21^. We therefore introduced an approach that makes use of the spatial information from the navigator read-out line to select data only within a small interval around the spinal cord. The first step was to calculate a 1D fast Fourier transform (FFT) of each navigator profile^9,20^, yielding a projection line along the frequency encoding direction.

**Figure 2:**
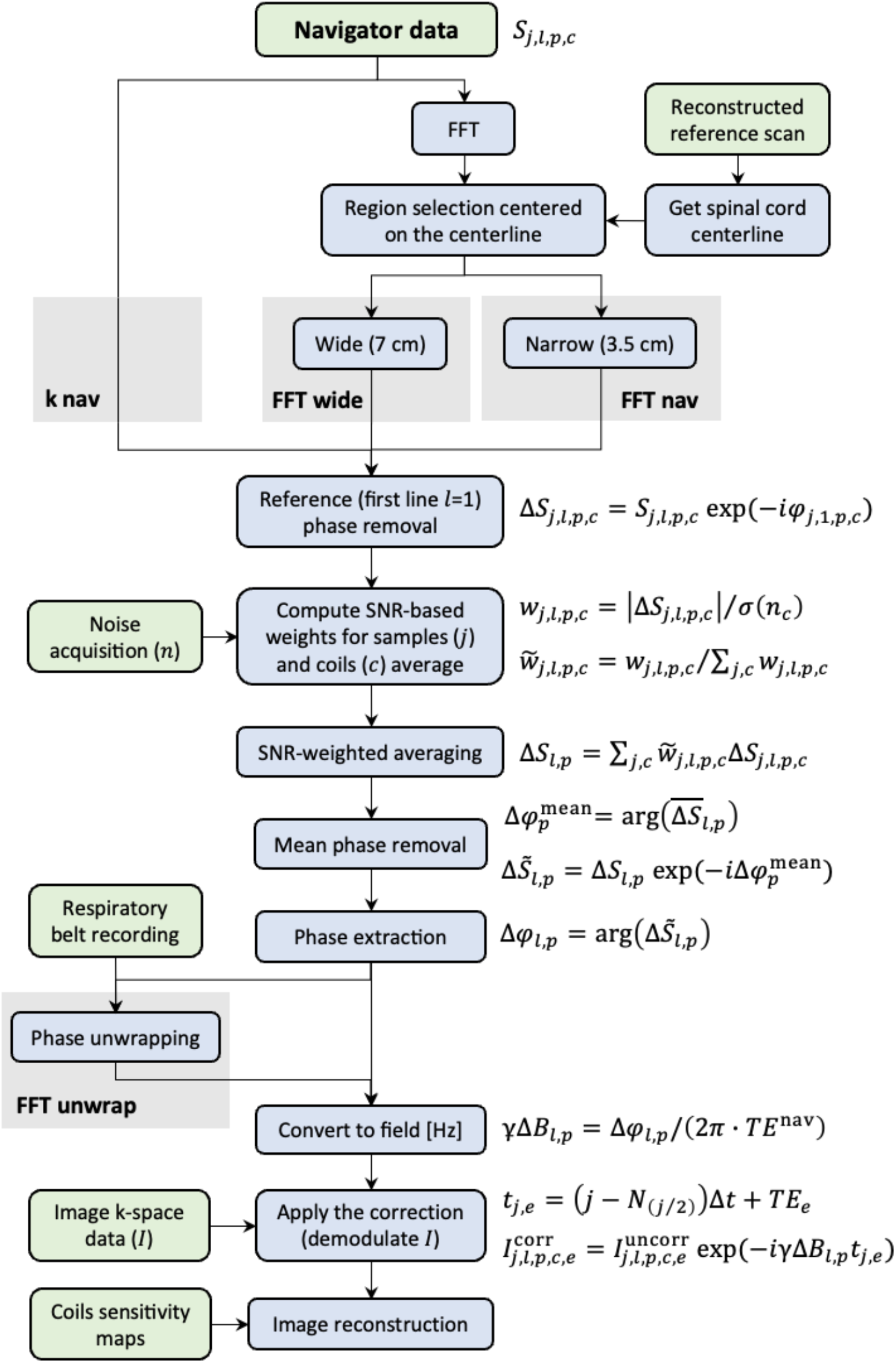
Processing pipelines for navigator-based corrections in the spinal cord. The required data and computation steps are represented by light green and light blue boxes, respectively. Four different approaches (grey background) were used to process the navigator profiles: (i) an optimized k-space based processing (*k nav*); two fast Fourier transform (FFT) based approaches, including region selection centered on the spinal cord centerline and covering (ii) a region of 3 cm approximately corresponding to the spinal canal (*FFT nav*) and (iii) a region of 7 cm corresponding to the vertebra (*FFT wide*); and (iv) an *FFT nav* processing with an added phase unwrapping step based on the respiratory belt recording data (*FFT unwrap*). Images were reconstructed after applying the navigator correction on the k-space image data by demodulating the signal. Each point in the k-space image data is identified by five indices representing the sample (j), line (l), slice (p), coil (c), and echo (e).

Subsequently, an interval of interest targeting the spinal cord was defined, discarding all samples outside the interval. The center of the interval was obtained by locating the spinal cord using the ‘sct_get_centerline’^31^ function in Spinal Cord Toolbox^32^ on the reference scan. When the automatic implementation of ‘sct_get_centerline’ failed, the manual selection method was used. Two different interval widths were considered for comparison: one of 3.5 cm, covering approximately the spinal canal (*FFT nav*), and a wider one of 7 cm (*FFT wide*), covering most of the vertebrae^33^ (Figure S1).

The following processing steps to extract phase estimates and apply the raw data correction were performed identically for the k-space navigator lines (*k nav)* and the projection lines (*FFT nav*/*FFT wide)*. To remove static phase contributions (static B_0_, B1^+/-^), the first navigator readout was chosen as reference, and its phase values (φ) were subtracted from all subsequent lines:

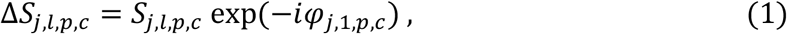

where *S*_*j,l,p,c*_ represents the navigator data and *j, l, p, c* are indices counting sample points, lines, slices, and coils, respectively. Within each slice, there was a large variability in signal contribution from different receiver coils and different sample points. Therefore, to maximize the SNR of the navigator data before phase extraction, an SNR-weighted averaging over samples and coils was performed:

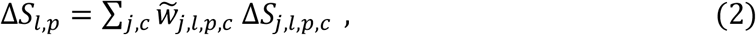

yielding one complex-valued data point per navigator line. The weights were computed as:

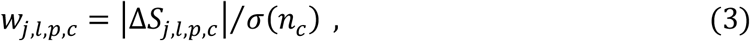

where *σ*(*n*_*c*_) was calculated as the standard deviation of the noise acquisition for each coil. The weights were normalized before use:

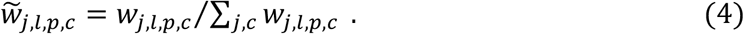

To reduce phase wrapping, we aimed to center the phase around zero before phase extraction. To this end, the phase of the mean navigator signal across lines (Δφ^mean^) was calculated:

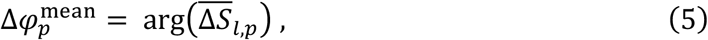

and subsequently subtracted from all profiles:

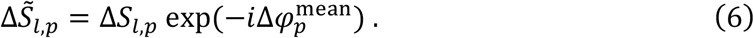

Only at this point were the navigator phase estimates calculated:

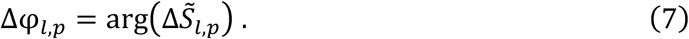

The phase estimates were then divided by the TE of the navigator readout (*TE*^nav^) to obtain field estimates scaled in Hz:

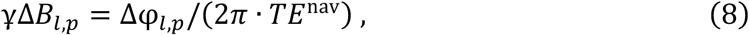

where γ is the reduced gyromagnetic ratio. The field estimates were used to calculate the expected phase variation at the time of each sample point in the image acquisition. The sample timing was computed as:

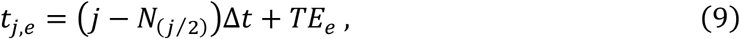

where *N*_*j*_ is the number of samples per line, Δ*t* is the time interval between sample points and *e* counts the echoes. Finally, the correction was applied to the raw data profiles from the imaging readouts 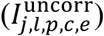 by demodulating the field fluctuations^19^, before the image reconstruction:

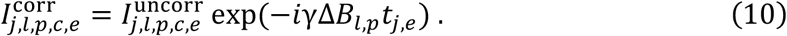

Occasionally, the extracted phase values were wrapped, yielding erroneous field estimates in Eq. 8. To address this, a phase unwrapping algorithm making use of the respiratory belt recordings was developed (*FFT unwrap*) and applied to the *FFT nav* approach. Details of the algorithm are given in the supplementary material. In brief, the belt data and navigator phase estimates were aligned by computing the cross-correlation peak after temporal smoothing. If a negative correlation was observed, the navigator estimates were inverted. Then, the navigator points corresponding to local minima in the belt data were averaged to define a baseline value, corresponding to complete expiration, for the navigator. Points near the local maxima in the belt data and below the baseline in the navigator estimates were identified as wrapped and corrected accordingly.

### Image reconstruction

Images were reconstructed without correction (*no nav*) and with correction based on the four different navigator processing pipelines (Figure 2). The image reconstruction was performed in MRIReco using a conjugate-gradient SENSE algorithm^30,34^ (10 iterations, L2 regularization). The sensitivity maps for the SENSE reconstruction were calculated by running the MRIReco implementation of the ESPiRIT^35^ algorithm on the reference acquisition, without masking. Masks for the sensitivity maps and the final reconstructions were calculated based on the reference images, as described in the supplementary material. A noise pre-whitening step, aimed at reducing the noise correlation across coils was applied before the reconstruction for SNR optimization purposes^36^. After reconstruction, the root-mean-square (RMS) image over the four echoes was computed. Correction effectiveness was evaluated both on the single-echo images and the RMS images.

### Quantitative analysis

To quantify the amplitude of the breathing-induced B_0_ field fluctuations along vertebral levels, the temporal standard deviation of the navigator field estimates from the *FFT unwrap* pipeline was calculated for each vertebral level and subject and was then averaged across repetitions. The amplitude sign was determined by the sign of the correlation between the navigator estimates and the belt recording (positive when in phase). To model the profile of the field fluctuations across subjects, an Eilers smoothing^37^ with 95% confidence interval was computed.

Multiple image quality metrics were computed to evaluate the performance of navigator correction. GM, WM, CSF and vertebral bodies (VB) were manually segmented, as described previously^24^, in JIM 7.0 (Xinapse systems, http://www.xinapse.com) on the RMS images averaged over repetitions from the *FFT nav* reconstruction (Figure S2). In the lumbosacral cord, only the most rostral vertebral body was segmented. The SNR was computed for GM and WM as

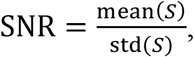

where *S* is the signal within the tissue of interest. The contrast-to-noise ratio (CNR) was computed between WM and CSF and between GM and WM as

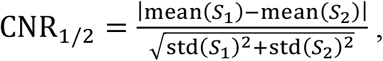

where the subscripts 1 and 2 indicate tissue types. The ratio between the mean signal in the CSF and the VB was also computed. This metric was designed to reflect the amount of ghosting artifacts. CSF and VB exhibit the highest and lowest signals in the image, respectively. Therefore, any ghosting is likely to decrease the signal in the CSF and increase the signal in the VB. The ratio was taken to eliminate general scaling factors. One advantage of this metric is that it does not rely on the standard deviation, which can be affected by factors causing signal variability within tissues, such as tissue inhomogeneity, static field inhomogeneity, bias fields, etc. In the lumbosacral cord, the CSF/VB ratio was calculated for only one slice pair located at the most rostral vertebra.

The quality metrics were calculated for the fourth echo and the RMS images of each repetition. The calculation was performed on a slice-by-slice basis and then averaged across two neighboring slices. In the cervical and thoracic cord, metrics were thus obtained for each slab (i.e., vertebral level). In the lumbosacral cord, we instead used two neuroanatomical landmarks^24,26^: the lumbosacral enlargement defined as the slice with the largest spinal cord cross-sectional area and the conus medullaris defined as the most caudal slice where the GM still has the characteristic butterfly shape. Four pairs of slices were considered: (i) the lumbosacral enlargement slice and the slice above (LSE), (ii) the two slices rostral to LSE (LSE+1), (iii) the two slices caudal to LSE (LSE-1), and (iv) the conus medullaris slice and the slice above (CM). In the final step, the quality metrics were averaged across the three repetitions.

## Results

### Field fluctuations

Figure 3 shows respiratory belt recordings and navigator estimates for a representative subject (subject 3) at all vertebral levels. The magnitude and polarity of the field fluctuations varied along vertebral levels but remained strongly correlated/anticorrelated with the respiratory trace. The profile of the field fluctuations along vertebral levels showed a similar trend across all subjects (Figure 4). In the upper cervical cord (C2-C4), the field magnitude was low and positively correlated with the belt recordings. The magnitude increased towards lower vertebral levels, peaking around C7 (5.0 ± 2.7 Hz). It then decreased towards a polarity inversion point around T3-T4. Below that point, the correlation was negative, with a mostly flat profile between T5-T7, then increasing slightly in magnitude towards T8 (−4.5 ± 1.7 Hz). Data were not acquired between T8-T12, but smoothing^37^ indicates the likely presence of a magnitude peak around T9-T10, with another polarity inversion around T11-T12. In the lumbosacral cord, the correlation was positive, and the magnitude reached a peak between T12 and L1 (5.1 ± 0.6 Hz).

**Figure 3:**
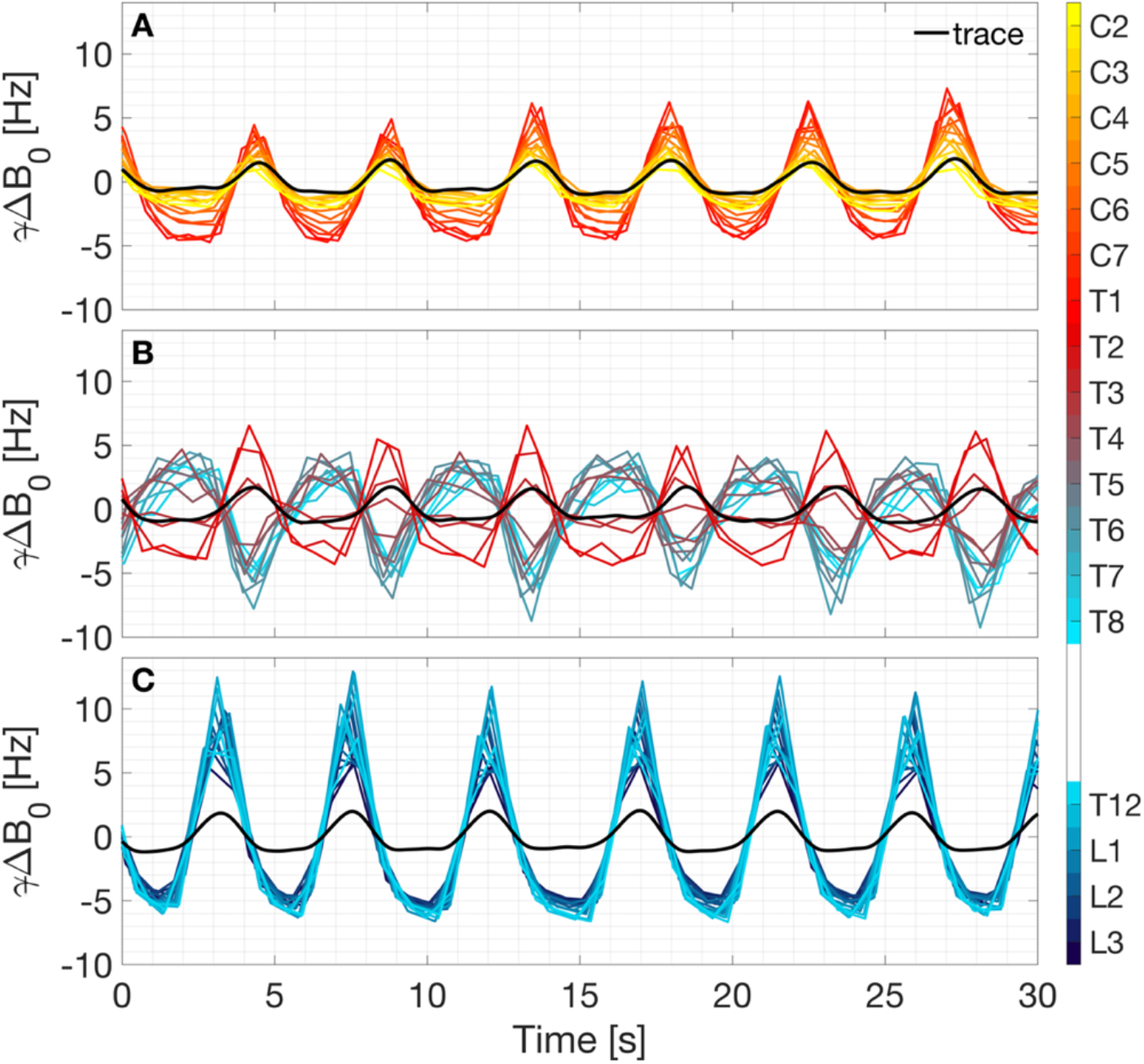
Navigator estimates for the breathing-induced field fluctuations, obtained with *FFT unwrap* navigator processing in subject 3 (first repetition), at the cervical (panel A), thoracic (panel B), and lumbar (panel C) vertebral levels. The respiratory belt trace recording (black line) is overlaid after filtering and alignment with the navigator traces, as described in the unwrapping algorithm (in the supplementary material).

**Figure 4:**
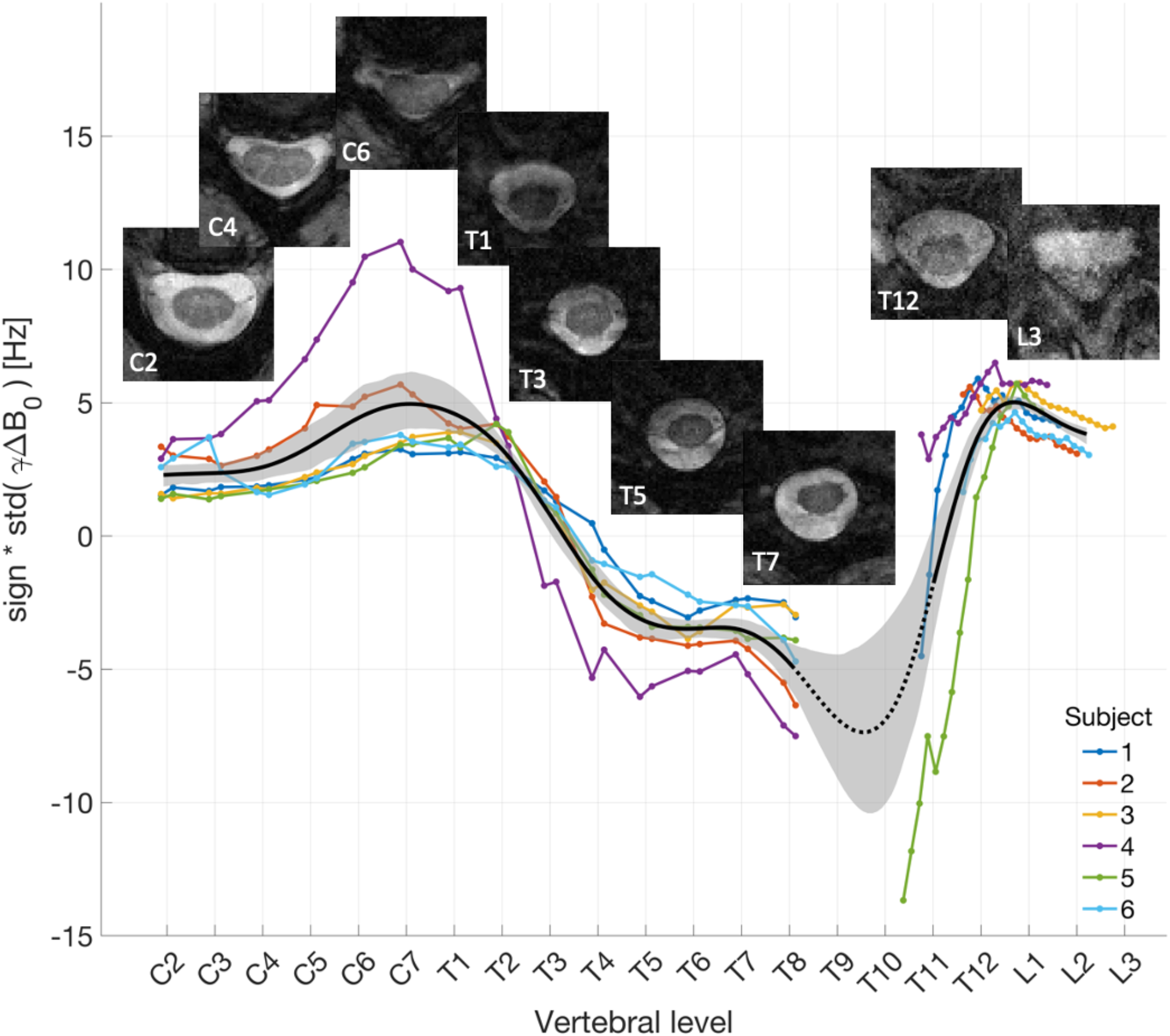
Profile of the breathing-induced B_0_ field fluctuations across vertebral levels. The magnitude represents the temporal standard deviation of the navigator field estimates from the *FFT unwrap* navigator processing pipeline. The sign was determined by the sign of the correlation between the navigator estimates and the belt recording (positive when in phase). To model the profile of the field fluctuations across subjects, an Eilers smoothing^37^ with 95% confidence interval was applied (black line and grey area). Data were not acquired between T8-T12, but the smoothing suggests the presence of a negative peak in this region (black dotted line). Images (TE=19 ms) without navigator correction are also displayed for subject 3.

### Qualitative image evaluation

The breathing-induced field fluctuations caused incoherent ghosting in images reconstructed without navigator correction. Vertebral levels with larger field fluctuations showed higher artifact load (Figure 4). Navigator-based corrections visually reduced ghosting artifacts in most slices and yielded more uniform signal within tissues, higher contrast between tissues, clearer anatomical details, brighter CSF, darker signal in the vertebral bodies, and reduced overall noise (Figure 5). The correction effect was more evident in slices with strong artifacts, often in the thoracic and lumbosacral cord (Figure 5) and at longer TE (Figure 6). Averaging across echoes and across repetitions (mean (RMS)) reduced the visual appearance of ghosting also without navigator correction. Nevertheless, the artifact load was visibly reduced by the navigator correction even in these cases.

**Figure 5:**
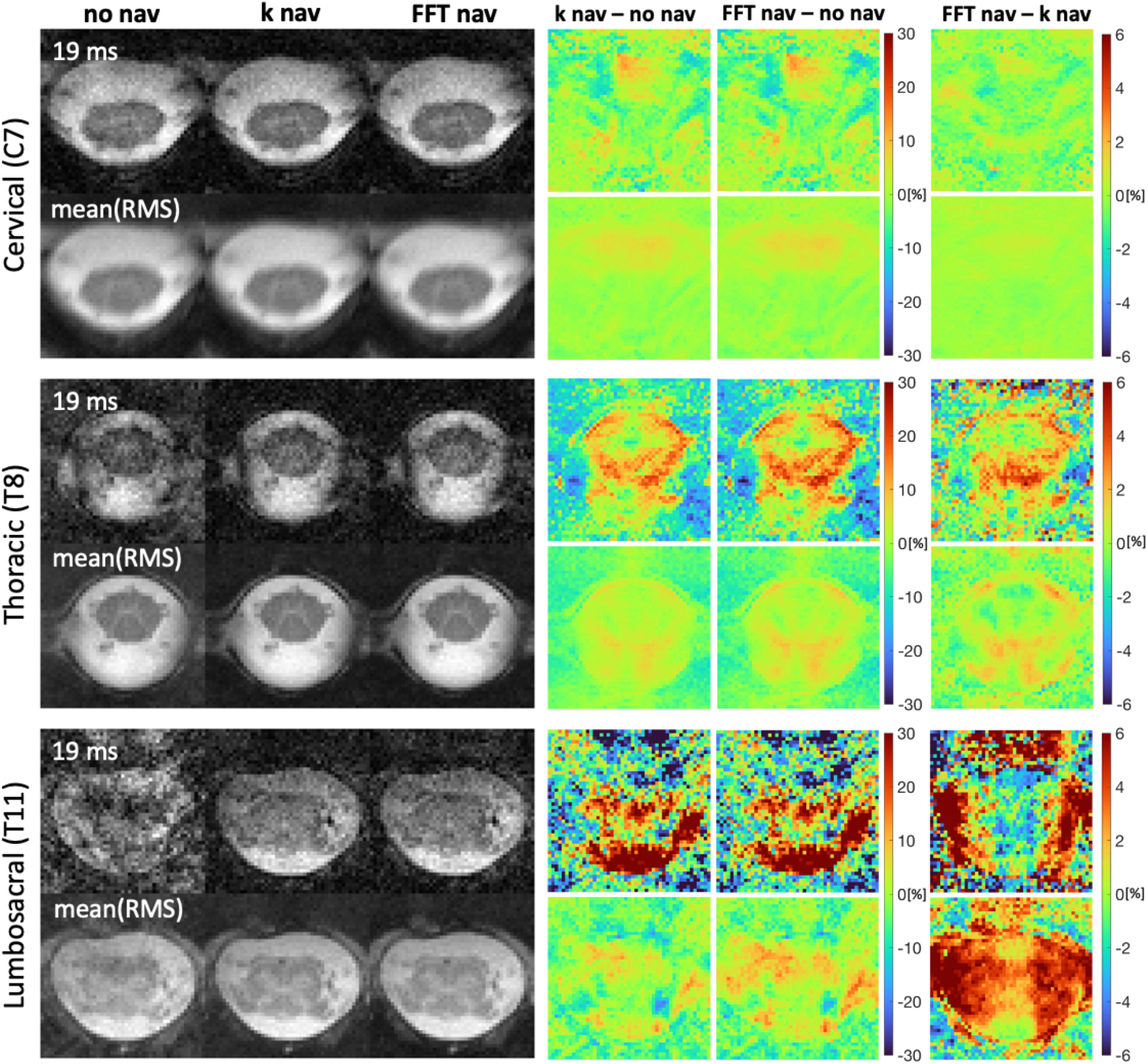
Images obtained in subject 5 by no navigator correction (*no nav*), *k* nav, and FFT navigator correction (*FFT nav*) for vertebral levels C7, T8, and T11. Displayed are a single repetition of the fourth echo (TE=19 ms), and the root-mean-square (RMS) images averaged across repetitions (mean(RMS)). Pairwise differences between the different navigator pipelines were computed after normalization (dividing each image by the maximum intensity value in that image).

**Figure 6:**
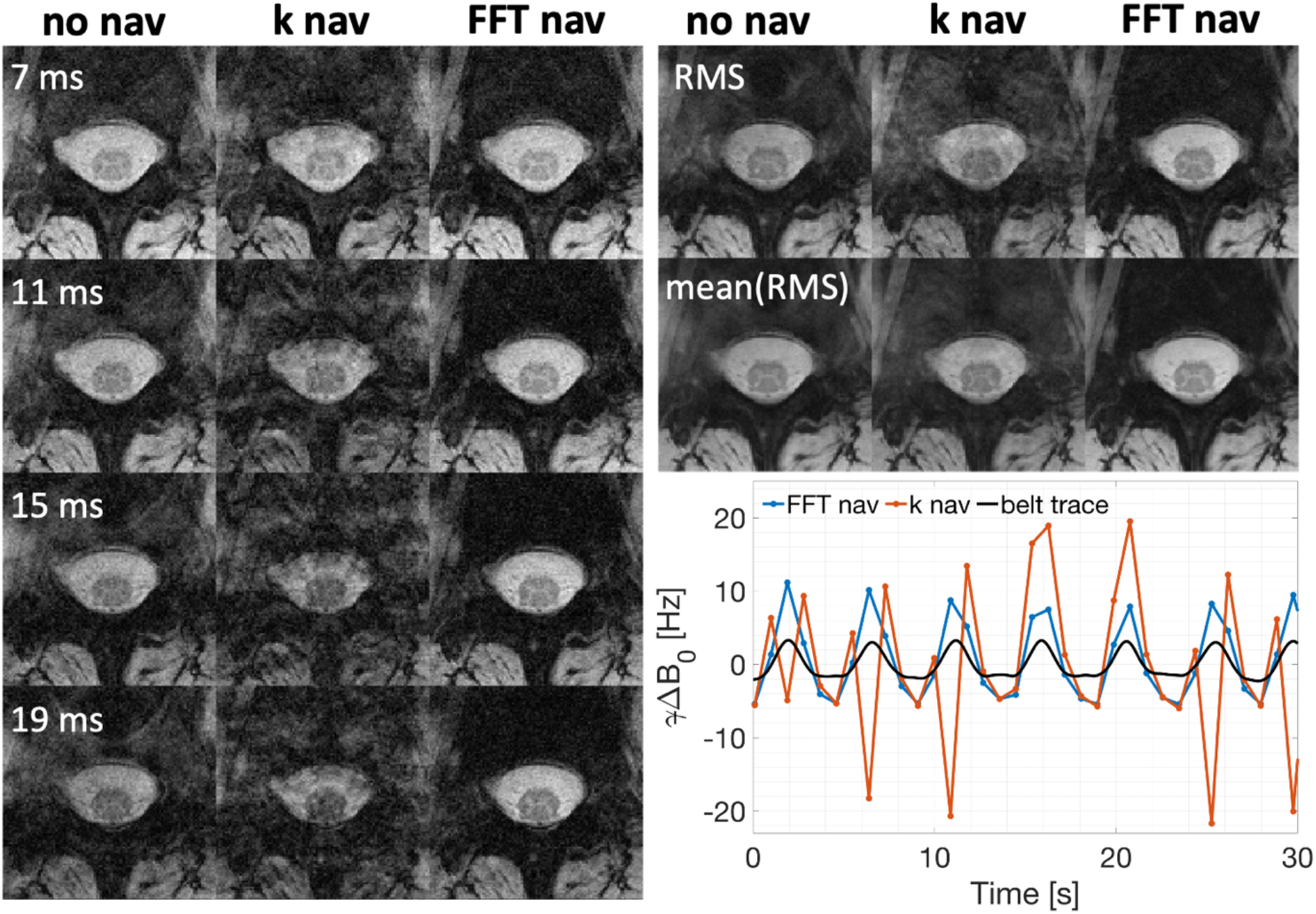
Images of a single slice positioned mid-T12 in subject 3, obtained after applying no navigator correction (*no nav*), *k nav*, and the FFT navigator correction (*FFT nav*). Displayed are a single repetition of all four echoes, along with root-mean-square (RMS) images for a single repetition and averaged across repetitions (mean(RMS)). The corresponding navigator field estimates are also shown in the bottom right.

Differences between images reconstructed without correction (*no nav*) and with correction (*k nav, FFT nav)* show that more signal is retained within the spinal cord and CSF when using navigator correction (Figure 5). In most cases, the difference between images resulting from different navigator pipelines was considerably lower than the difference between corrected and uncorrected images. In some cases, however, *k nav* yielded irregular phase estimates, and exacerbated ghosting compared to no correction (*no nav*) (Figure 6). The *FFT nav* generally performed well also in these cases, and never reduced the image quality compared to *no nav*. FFT navigator corrections based on two different spatial intervals (*FFT nav*, and *FFT wide*) yielded visually indistinguishable results in most cases. When a visual difference was apparent, the narrower interval yielded better image quality in most, but not all, cases (Figure 7).

**Figure 7:**
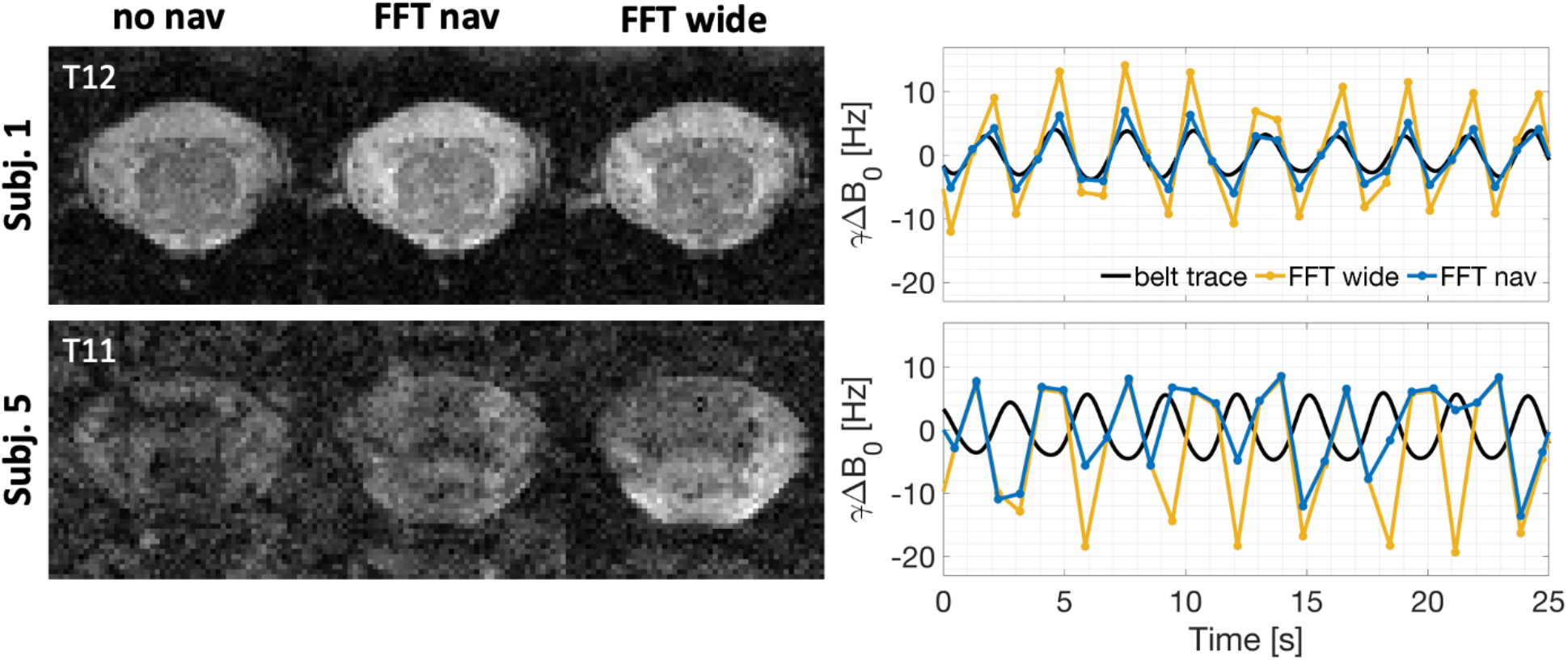
Comparison of reconstructed images (TE=19ms) and navigator field estimates for different sizes of the region selection interval in the FFT approach. For *FFT nav* an interval size of 3.5 cm was used, approximately corresponding to the spinal canal size. For the *FFT wide* case, this was set to 7 cm, approximately matching the size of the vertebral body. The upper row (subject 1) illustrates the more common scenario where a narrower interval selection results in improved image quality compared to the wider interval. The lower row (subject 5) displays one case where the wider interval performed better.

In most slices, the navigator-based field estimates were not affected by phase wrapping. Wrapping occurred in a few slices of the lumbosacral acquisitions, predominantly in subjects 1 and 5. The affected slices were at the level of lower thoracic and upper lumbar vertebrae, close to the lower edge of the lungs. In these cases, an unwrapping step (*FFT unwrap*) was necessary to accurately capture the field fluctuations (Figure 8). Only navigator points collected close to peak inspiration, corresponding to the peak field fluctuations, were wrapped. In rare cases, when subjects took a deep breath, one or two points were wrapped in otherwise unaffected slices. *FFT unwrap* successfully corrected most, but not all, of the wrapped points and yielded visually improved image quality compared to *FFT nav* (Figure 8). In slices without wrapping, *FFT unwrap* yielded field estimates identical to *FFT nav* for most points. In rare cases, a few points were incorrectly identified as wrapped, but this did not visually affect the image quality. The unwrapping algorithm used data from several slices for the synchronization, and information from all data points within one slice to identify wrapped points. Its performance could therefore be influenced by the TR of the acquisition, as the TR determines the sampling rate of the breathing cycle. However, *FFT unwrap* performed equally well for both TRs tested (Figure 8).

**Figure 8:**
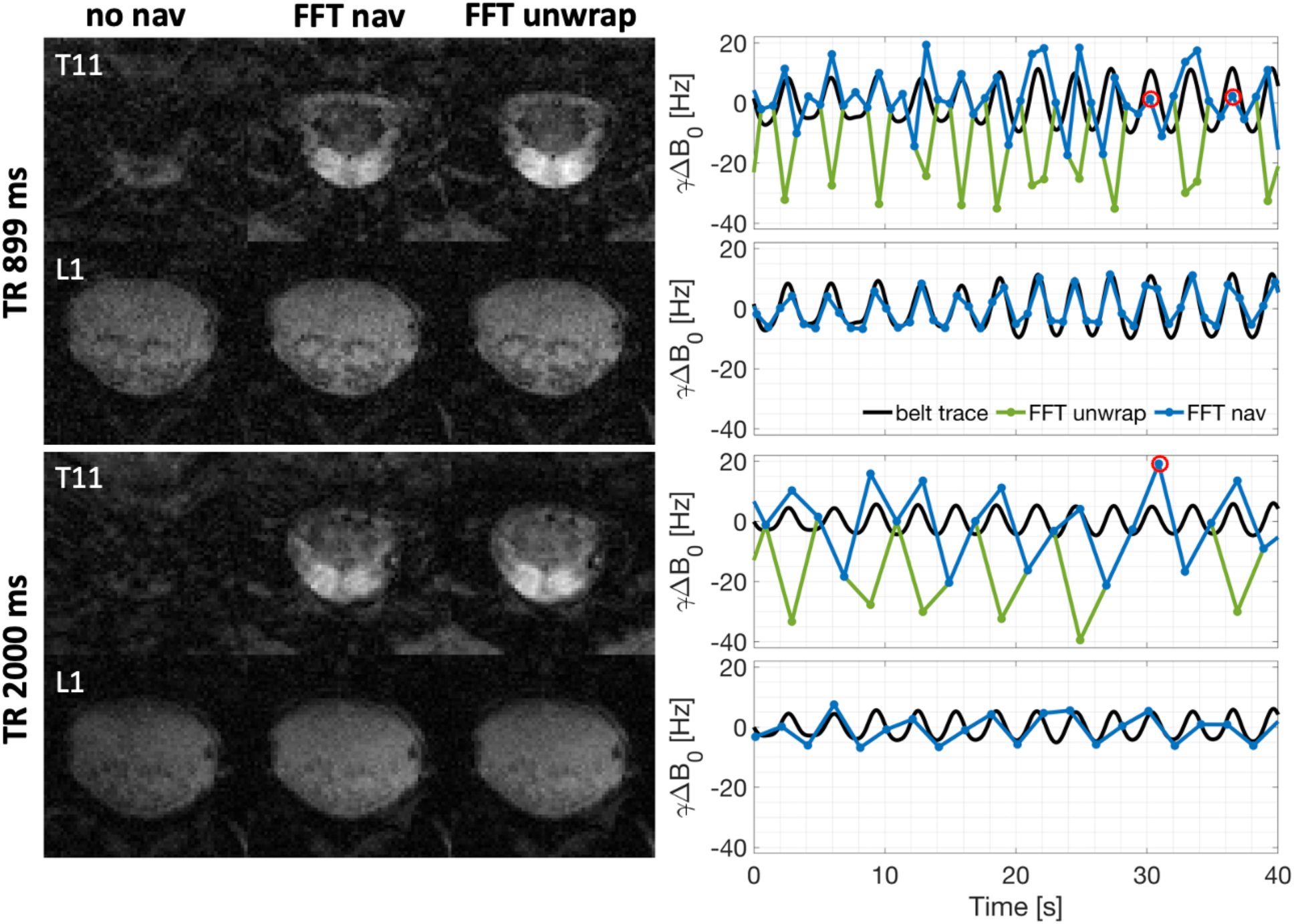
Comparison of reconstructed images (TE=19ms) and navigator field estimates for the FFT navigator with and without phase unwrapping in subject 5. Only the fourth echo images are displayed as they show the biggest correction effect, facilitating the comparison. Two TRs were considered (899 ms and 2000 ms) as the unwrapping algorithm is the only part of the navigator processing that may be TR dependent. For each TR, two slices are shown, one presenting heavy phase wrapping (at level T11) and one presenting no phase wrapping (at level L1). A few wrapped points were not corrected by the algorithm and are marked with red circles on the navigator traces.

### Quantitative image evaluation

Pairwise comparisons of quantitative metrics between *no nav, k nav*, and *FFT nav* are shown for the fourth echo (TE=19 ms) in Figures 9 and S4. Results for the RMS images were very similar but with smaller effect sizes (Figures S3, S5). All quantitative metrics exhibited an average improvement with both correction approaches relative to the *no nav* case. The improvement showed a pattern along the vertebral levels that roughly resembled the pattern of field fluctuations (see Figure 4). Thus, in agreement with the qualitative results, slices with higher artifact load before correction (lower cervical/upper thoracic and lower thoracic/upper lumbosacral cord) showed larger improvement in the quantitative metrics. The pattern was most pronounced in the CSF/VB ratio, which also appeared to be less noisy compared to the other metrics, indicating that CSF/VB ratio is a sensitive measure of artifact load.

**Figure 9:**
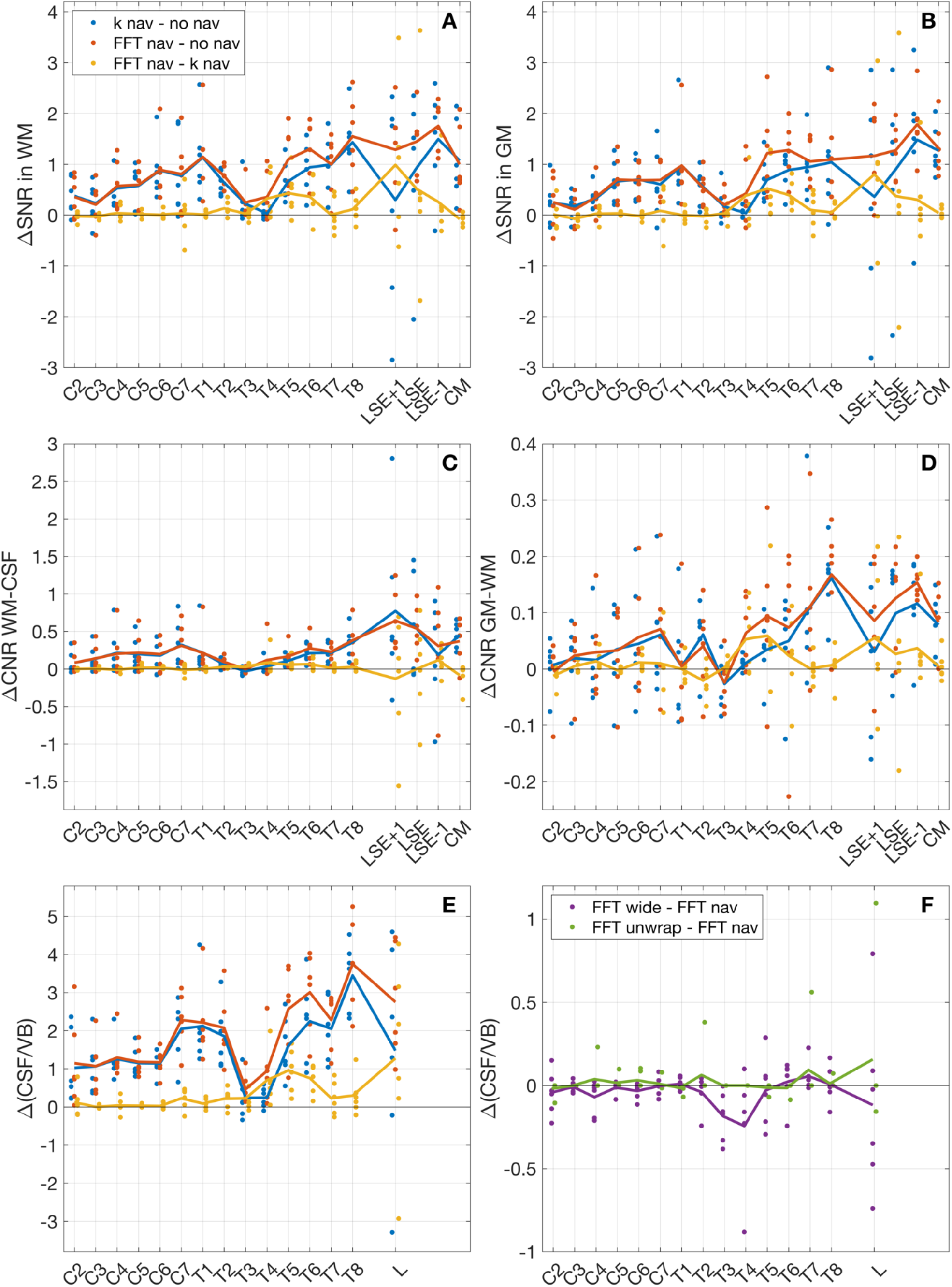
Comparison of image metrics between *no nav, k nav*, and *FFT nav* (panels A-E), and between *FFT unwrap, FFT wide*, and *FFT nav* (panel F) for the fourth echo (TE=19 ms). Each data point represents one pair-wise comparison for one subject and vertebral level. The solid lines connect the group-average image metrics across vertebral levels. Only the difference in CSF/VB signal ratio is shown for the comparison between the FFT pipelines, as this metric is the most sensitive to the artifact load.

The *k nav* and *FFT nav* corrections exhibited comparable improvements in the quantitative metrics in regions with low field fluctuations. In regions with large field fluctuations, especially in the thoracic and lumbosacral cord, there was a clear advantage for *FFT nav* (Figure 9). Pairwise comparisons between *FFT wide* and *FFT unwrap* vs. *FFT nav* are shown in Figure 9F for the CSF/VB ratio and in Figures S3-S5 for the other metrics. While the differences were small, *FFT wide* yielded equal or slightly worse results at most vertebral levels as compared to *FFT nav. FFT unwrap* slightly improved image quality metrics compared to *FFT nav* in regions with more intense field fluctuations, where phase wrapping is more likely to be present.

To assess the impact of the improvement in quantitative metrics, we here report the group-average percentage increase in SNR and CNR for *FFT nav* compared to *no nav* at the two locations with the largest field fluctuations (Figure 4): C7-T1 and T8-LSE+1. The percentage increases were computed on the group-average metrics for each vertebral level, averaged across C7-T1 and T8-LSE+1, and are reported as (C7/T1 mean ± std, T8/LSE+1 mean ± std). For the fourth-echo images (TE=19 ms), the percentage increases were: SNR WM (22±21%, 29±28%), SNR GM (15±16%, 20±26%), CNR GM-WM (9±37%, 28±37%), CNR WM-CSF (19±29%, 37±41%). For the RMS images, the percentage increases were: SNR WM (15±14%, 16±15%), SNR GM (5±8%, 8±14%), CNR GM-WM (11±20%, 12±25%), CNR WM-CSF (9±11%, 15±20%).

Figure 10 shows the SNR, CNR, and CSF/VB ratio across vertebral levels for *FFT nav*. The SNR in GM and WM was approximately 50% higher in the RMS images than in the fourth echo. Additionally, the RMS images showed a slight CNR increase between GM and WM, as well as between WM and CSF. While the SNR and CNR values displayed a relatively flat profile across vertebral levels, the CSF/VB ratio somewhat resembled the pattern of field fluctuations in both the fourth echo and RMS images. This suggests that the CSF/VB ratio is a more sensitive measure of residual artifact load than SNR and CNR, in consistence with the results in the pairwise comparison. The CSF/VB values were higher in the fourth echo images than in the RMS images because of the long 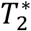 in CSF.

**Figure 10:**
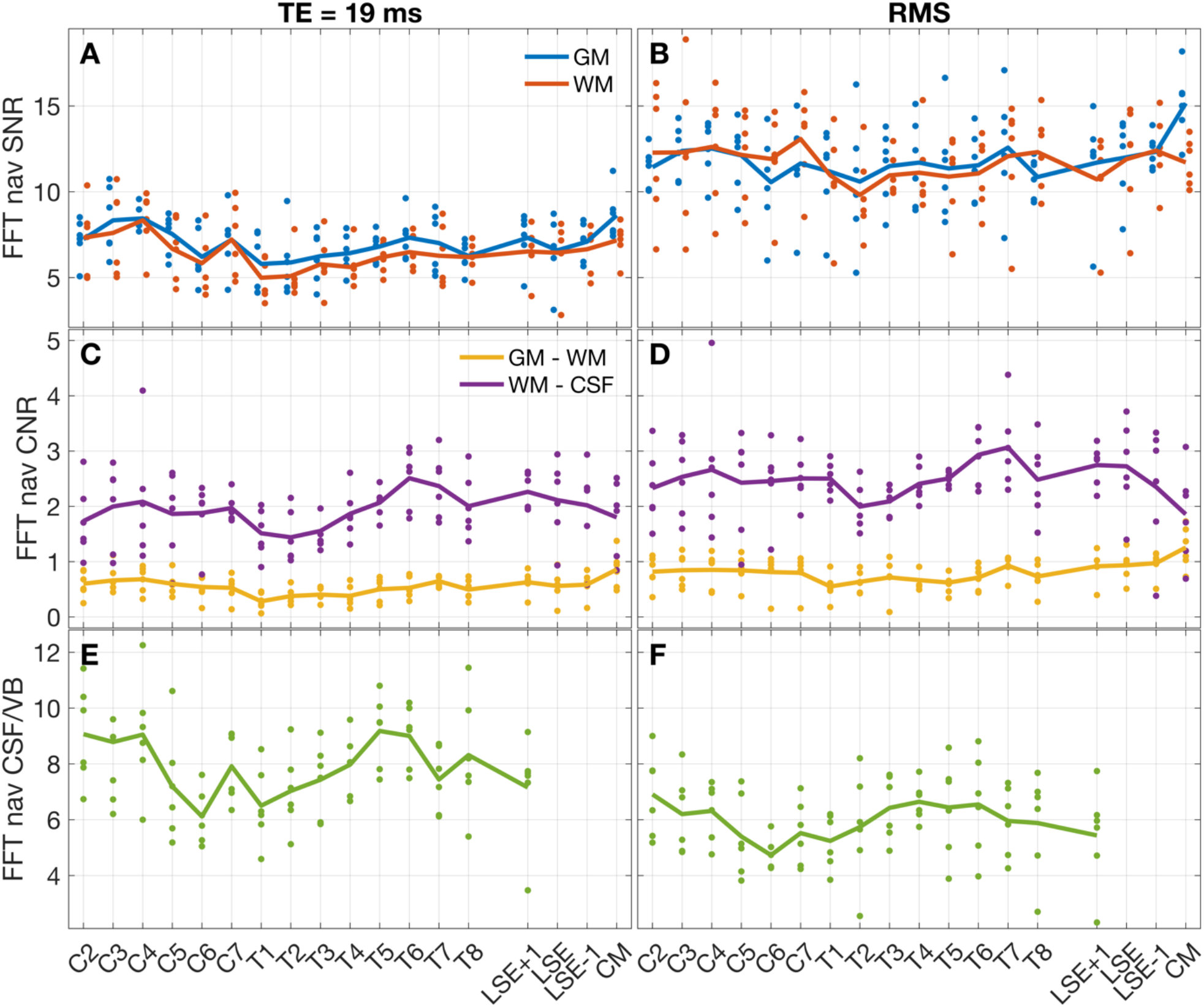
Mean SNR, mean CNR, and CSF/VB ratio across subjects, for each vertebral level. The values were computed on the fourth echo (TE = 19 ms) and on the root-mean-square (RMS) images obtained with the *FFT nav* pipeline and then averaged across repetitions.

## Discussion

The aim of this study was to achieve a robust and effective correction for breathing-induced B_0_ field fluctuations in the spinal cord. To make the correction easily transferable between sites, we focused on retrospective correction based on a common navigator, thus not requiring specialized hardware or extensive sequence modifications. The navigator data processing was designed to yield robust field estimates under the challenging conditions of spinal cord imaging. A few key steps were important for robustness: i) FFT to select a spatial interval, ii) SNR-weighted averaging over coils and samples, iii) averaging on the complex data to maximize SNR before phase extraction, iv) centering the phase to reduce wrapping.

While standard navigator processing frequently fails in the spinal cord, sometimes even exacerbating the artifacts, the optimized processing improved image quality in most cases and at worst yielded comparable images to the uncorrected case. The proposed navigator processing contains several configurations. We advise using the *FFT* processing, setting the spatial interval to approximately cover the spinal canal and adding the phase unwrapping step in case of confirmed phase wrapping in the navigator field estimates.

The *k nav* pipeline assumes spatially homogeneous field fluctuations in the whole slice, while the FFT pipelines reduce this assumption to the selected interval. The interval selection takes advantage of the ghosting pattern in line-by-line Cartesian imaging, where ghosting appears in the phase encoding (PE) direction only. Consequently, ghosting originating from outside the region of interest in the readout direction is of less concern. However, the approach is therefore highly dependent on the PE direction. AP phase encoding is the most practical choice in the lower cervical, thoracic, and lumbosacral spinal cord, but not necessarily in the upper cervical cord. The interval selection requires a choice of center and width of the interval. In this work, the center was set for each slice individually based on an automatic spinal cord segmentation algorithm^31^. Slice-wise identification of the spinal cord may be particularly relevant for subjects with anatomical variations, such as scoliosis. The width of the interval presents a trade-off between sensitivity and accuracy: while a wide interval yields more data points that can contribute to SNR, a narrow interval better fulfills the assumption of spatially homogeneous field fluctuations. Two interval widths were compared here, showing a tendency towards higher image quality with the narrower interval, which approximately corresponded to the spinal canal.

Phase wrapping can degrade the results of the navigator correction. Wrapping is more likely to occur in regions with strong field fluctuations, in acquisitions with long TE, and on high-field MR systems. The sampling rate of the breathing cycle in each slice depends on the TR of the acquisition and is often too low to rely on temporal phase unwrapping alone. We therefore added an optional unwrapping step, making use of external information from a respiratory belt. The performance of the proposed unwrapping was here evaluated in combination with the FFT pipeline, but the algorithm can also be applied to the *k nav* pipeline. We observed a clear trend towards improved image quality with the proposed unwrapping algorithm. However, the unwrapping occasionally failed, being more prone to failure in the presence of highly wrapped phase We advise visually assessing its performance and reverting to the original FFT in the case of irregular phase estimates.

The strongest image artifacts were observed close to the upper (C7-T1) and lower (T8-T11) borders of the lungs, in correspondence with peaks in the local field fluctuations. Many spinal cord MRI studies have focused on the upper cervical spinal cord, where the artifacts are less intense, with fewer studies showing data from the lumbosacral cord^24,38–40^. However, imaging of lower spinal cord levels is clinically important and highly relevant for neuroscientific investigations. The proposed navigator-based correction was shown to be effective in every spinal region, with the largest improvement in regions with higher artifact load. The improved image quality provides significant potential benefits throughout the spinal cord. It may allow to reduce the scan time, by decreasing the number of averages, and could potentially improve automatic segmentation outcomes. Most importantly, it may increase the reliability of imaging results. Even subtle ghosting artifacts can distort quantitative assessment, in particular when single echo-images are analyzed. Applying the optimized correction may improve the reproducibility and repeatability of quantitative results, which is especially important for longitudinal studies.

We used the navigators to characterize the profile of field fluctuations along vertebral levels. Such characterization may facilitate further development of prospective or retrospective correction techniques but has previously been performed in the cervical and upper-mid thoracic spine only^15,21^. The navigator-based field measurements yielded field estimates that are broadly consistent with previously published results. While all studies showed a similar spatial pattern in the cervical and upper thoracic spinal cord^15,21^, the magnitude of the field fluctuations was reported to be considerably larger in Verma et al (74 Hz at C7 at 3T).

Multiple experimental aspects could explain this inconsistency, including the depth of inspiration during the breath-holds as well as differences in the pool of volunteers.

The observed field pattern is consistent with a model of the lungs as air-filled spheres immersed in water, which expand upon inspiration^41^ (field simulation results in the supplementary materials, Figure S6). Because air has positive magnetic susceptibility relative to water, such a model predicts a positive field shift during inspiration in locations superior and inferior to the lungs and a negative field shift between the lungs. Indeed, in the central mid-thoracic cord, the navigator field estimates showed a negative correlation with the respiratory belt recording, while the correlation was positive in the cervical and lumbosacral cord. We did not have data around T8-L1, which corresponded to the lower sign inversion point in most subjects. Future work may characterize the field fluctuations in the full spinal cord. The characterization may however be affected by the subject positioning and the MR system model, as the tissue magnetization within the thorax depends on the distance to the isocenter, as well as the B_0_ field profile along the scanner bore.

While image quality was generally improved by the correction, there were residual ghosting artifacts, especially in regions with large field fluctuations. The correction may be further improved by going beyond the assumption of spatial homogeneity, to include linear gradients or even higher-order fields. The existing navigator data would allow for fitting field fluctuations along the LR readout direction. However, due to symmetry, the main field gradients are expected in the AP and FH directions^21^. In brain imaging, it has been proposed to extract spatial B_0_ field information from single lines navigators^10^ or FID navigators^42,43^ using the coil sensitivity profiles. The same approach may be applicable in the spinal cord^44^ but may require specific optimization due to different coil sensitivities and spatial field profiles. Alternatively, full 2D/3D navigators may be employed^8,45^, but they would take up more time in the acquisition. Instead of navigators, field probes can be used to monitor time-varying fields^46^, but would most likely require additional optimization for spinal cord applications. Dynamic gradients in the through-slice direction may additionally cause non-recoverable effects, such as time-variable signal dephasing, which cannot be addressed with retrospective correction. The through-slice field gradient was largest near the correlation sign inversion points, where the magnitude of the field fluctuations otherwise was low.

Addressing these effects would require prospective correction, such as real-time shimming^7,11,14,16,18^.

Correction of breathing-induced B_0_ field fluctuations was implemented for anatomical 2D ME-GRE imaging of the spinal cord at 3T. However, many other applications may also benefit from a similar approach. Breathing-induced B_0_ field fluctuations are even stronger at higher background field strengths, and effective correction becomes even more important. In the case of 3D acquisitions, the ghosting will likely be less distinct due to the averaging effect of the additional phase encoding steps. However, the field fluctuations are still likely to affect the SNR of the images. 3D acquisitions may require the inclusion of linear gradients for effective correction, due to the larger slab thickness. Beyond anatomical ME-GRE, segmented EPI acquisitions for fMRI or diffusion-weighted imaging will also be vulnerable to field fluctuations between segments and would benefit from similar correction. Finally, the pipeline for robust phase estimation, including spatial region selection, may also be relevant for other anatomies where the region of interest constitutes a small part of the full FOV.

## Conclusion

In this study, we proposed optimized processing pipelines for robust navigator-based retrospective correction of breathing-induced B_0_ field fluctuations in the spinal cord. We demonstrated that the correction reduced ghosting artifacts, thereby improving image quality and increasing SNR and CNR, in anatomical ME-GRE acquisitions in all regions of the spinal cord. The enhanced image quality holds promise to improve the diagnostic value of ME-GRE imaging and increase the reliability of quantitative analyses. The ease of implementation across sites makes the technique attractive for clinical and scientific applications.

## Acknowledgments

We thank all the volunteers who participated in this study. We also thank Veronika Birkhäuser and Oliver Gross, as well as the entire clinical team of the Department of Neuro-Urology, Balgrist University Hospital, University of Zürich for screening the healthy volunteers. We would further like to express our appreciation to Lars Kasper (Toronto NeuroImaging Facility (ToNI), Department of Psychology, University of Toronto), for guiding our initial work with the MRIreco.jl package. This work was financially supported by the Swiss National Science Foundation (SNSF) (33IC30_179644). Imaging was performed with the support of the Swiss Center for Musculoskeletal Imaging, SCMI, Balgrist Campus AG, Zürich.

## Data availability statement

All proposed navigator processing pipelines are collected in a Julia package called MRINavigator.jl v0.1.1 (https://github.com/NordicMRspine/MRINavigator.jl). Documentation is available online (https://nordicmrspine.github.io/MRINavigator.jl/dev/) along with example data acquired on a phantom (https://doi.org/10.5281/zenodo.10731729), example scripts and expected outcomes (https://github.com/NordicMRspine/UserExample_MRINavigator/tree/main).

## Notes

### Competing Interest Statement

The authors have declared no competing interest.

### Funding Statement

This study received funding from the Swiss National Science Foundation (SNSF) (33IC30_179644). Imaging was performed with the support of the Swiss Center for Musculoskeletal Imaging, SCMI, Balgrist Campus AG, Zürich.

### Author Declarations

Ethics committee of Kantonale Ethikkommission Zürich, BASEC ID: 2019-00074 gave ethical approval for this work

