## Supplementary material for "Optimized navigator-based correction of breathing-induced B_0_ field fluctuations in multi-echo gradient-echo imaging of the spinal cord"

**Outline:**

- Methods (page 2)
    - Figures S1, S2
- Results (page 4)
    - Figures S3 - S5
- Discussion (page 7)
    - Figure S6
- References (Page 9)

#### Methods

##### Masking the coil sensitivities

To obtain masks for the sensitivity maps, the reference acquisition was reconstructed without SENSE, and the root-sum-squared (RSS) image across coils was computed. A rough mask for each slice was initially obtained by defining a threshold on the RSS image (13% of the maximum) and applying it to each slice individually. The biggest connected component for each slice was identified, and voxels outside it were removed from the mask. Breathing-induced ghosting artifacts in the reference images sometimes led to voxels located posteriorly to the volunteer's back being included in the mask. To identify the posterior border of the back, the mask was projected in the AP direction and the projection derivative was computed. The border was defined by computing the maximum of the derivative in the posterior half of the image. A margin of 3 voxels posterior to the identified border was applied, and all voxels posterior to this line were removed from the mask. Finally, to make the mask uniform, all voxels inside its convex hull were included.

##### Phase unwrapping algorithm

The first algorithm step consisted of smoothing the respiratory belt data with a Butterworth low-pass filter (cut-off frequency 0.7Hz, 3 poles). Low-frequency oscillations were removed from the navigator phase data and the belt data by applying a Butterworth high-pass filter (cut-off frequency 0.15Hz, 5 poles). An additional alignment step between the traces was needed, as a delay between the respiratory movement and the field fluctuations was noticed in most subjects. Not all the navigator slices were considered for the alignment as some presented noisy traces or prominent phase wrapping. The correlation between the respiratory and navigator traces was computed and only the navigator slices with a correlation within $1.7\sigma$ from the maximum correlation in the acquisition were considered for the alignment. The selected navigator profiles were combined in a single vector and low-pass smoothed. In the case of a negative correlation[1], the navigator estimates were inverted before the combination. The time shift was estimated by locating the peak of the cross-correlation of the belt recordings and the so-built navigator vector. After the alignment, the shifted belt recordings were interpolated to the navigator time points for each slice. The correlation between the belt and navigator traces was re-computed and the navigator estimates were inverted in case of negative correlation. As slices with strong wrapping can give negative correlation values, only up to one sign change was allowed across slices in the same acquisition. Its position was found smoothing the correlation vector (low-pass filter cut-off frequency 0.075Hz, 3 poles). Slices in which the navigator phase estimates $2\sigma$ interval was lower than $0.9\pi$ were considered unlikely to include wrapped points and excluded from further analysis. Also slices with a correlation lower than 0.2 were excluded, to prevent an inaccurate respiratory trace from misleading the unwrapping. At this stage, the wrapped points could be identified. To do that, the navigator baseline value, corresponding to the complete expiration phase was computed for each slice. This was obtained as the average of the navigator phase values corresponding to the lower 20% intensity points in the respiratory trace. The baseline was subtracted from each slice's navigator trace. The wrapped points were those in the top 30% of the belt trace and below $-0.6\pi$ in the navigator trace. The numerical parameters in the algorithm were tuned empirically, by testing multiple values and visually investigating their effect on the traces. The parameter tuning was performed on a previous set of lumbosacral data consisting of ten healthy subjects. Six of the ten subjects also volunteered to be scanned for this study.

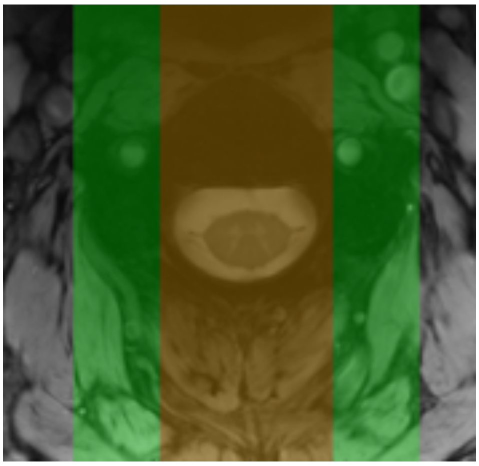

**Figure S1:** Two region selection interval sizes were used and compared for the FFT approach. The smaller one (in red) with a width of 3.5 cm, covering approximately the spinal canal. The second one (in green) with a width of 7 cm, covering most of the vertebrae.

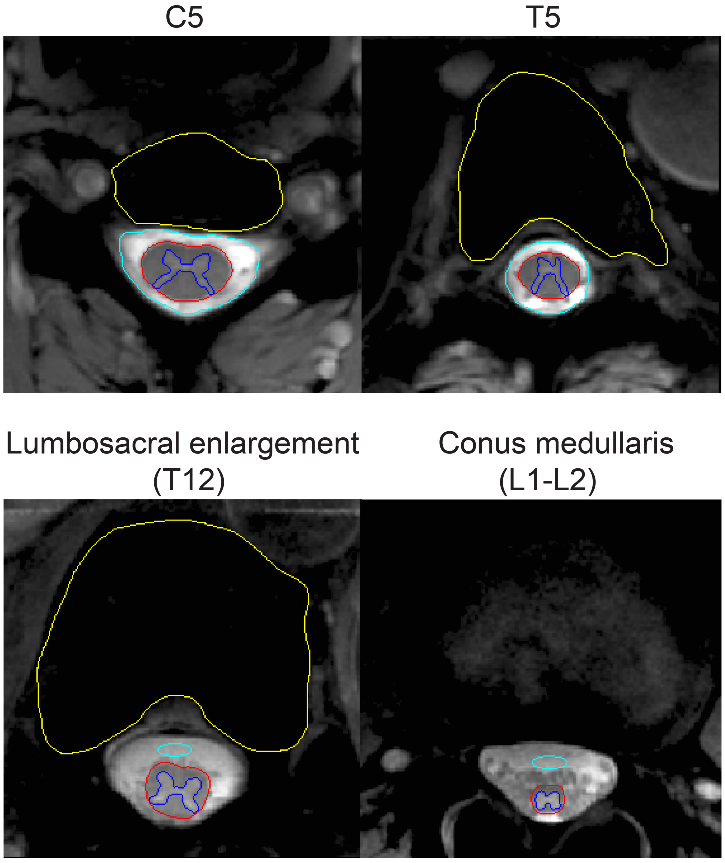


**Figure S2:** Example of grey matter (blue outline), white matter (red outline), cerebrospinal fluid (light blue outline), and vertebral body (yellow outline) segmentation at multiple vertebral levels (C5, T5, T12, L1-L2) on a representative subject.

### Results

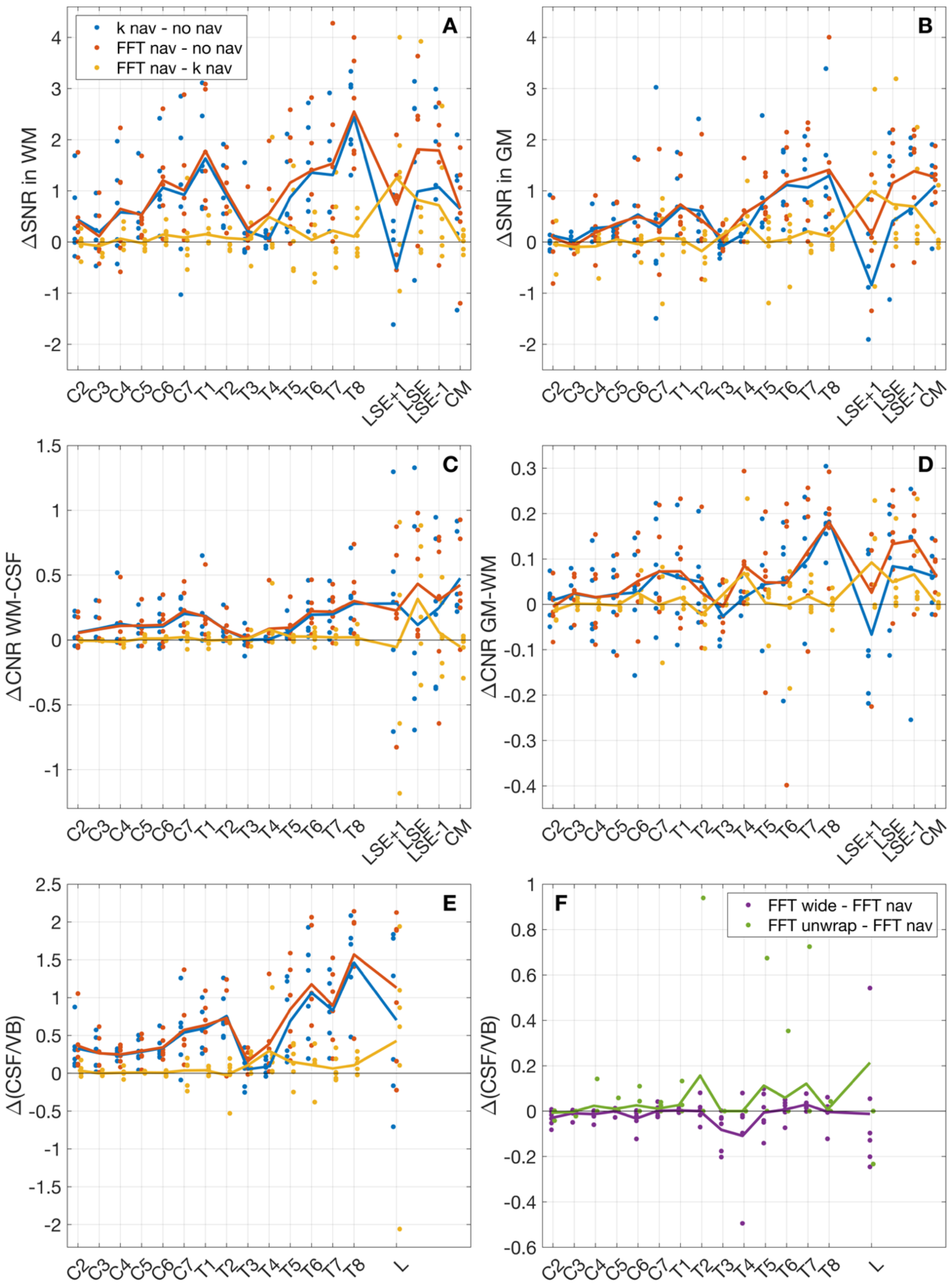


**Figure S3:** Comparison of image metrics between *no nav*, *k nav,* and *FFT nav* (panels A-E), and between *FFT unwrap, FFT wide,* and *FFT nav* (panel F) for the root-mean-square (RMS) images. This figure complements Figure 9 in the main text. Further details can be found in that figure caption.

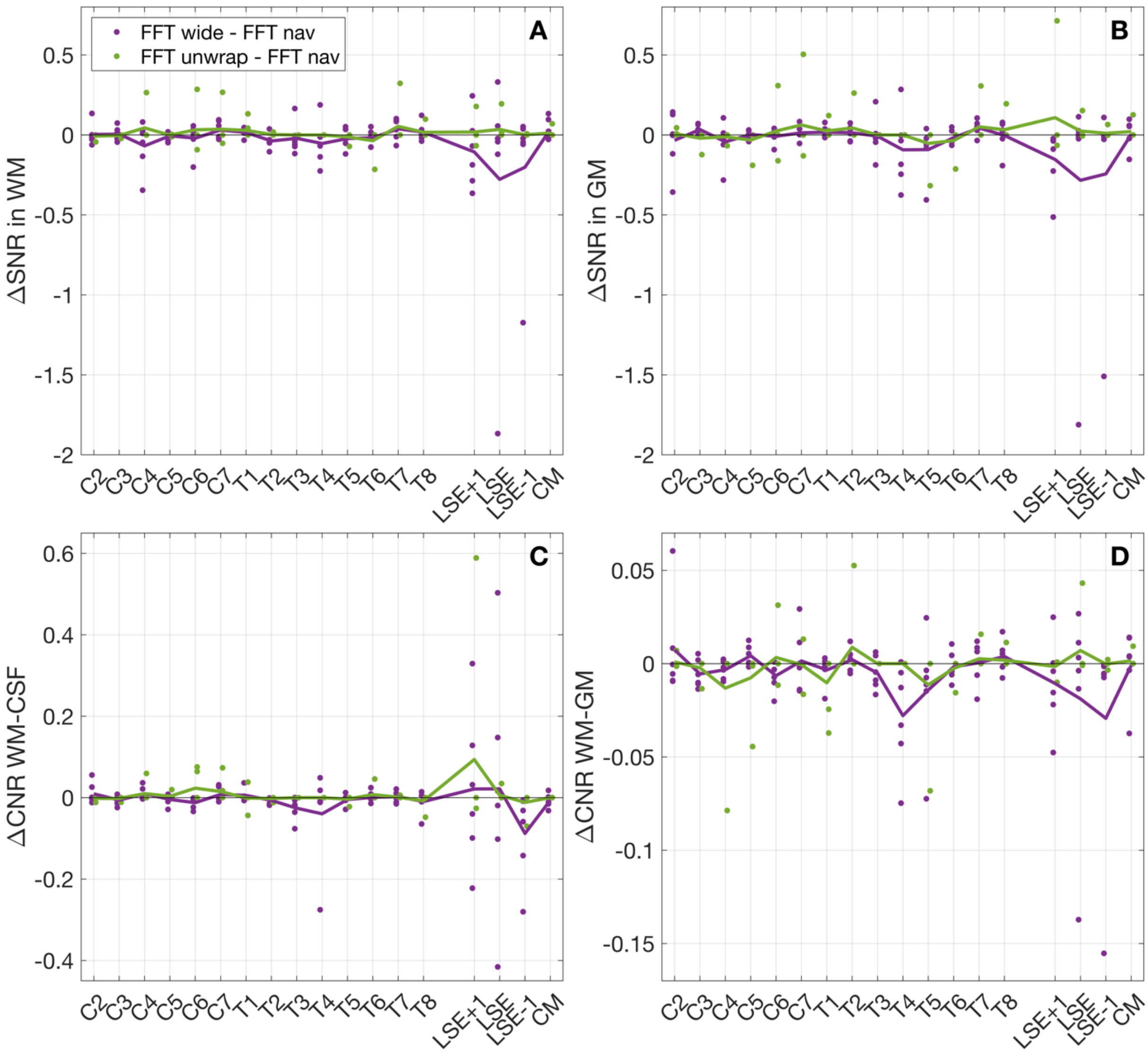


**Figure S4:** Comparison of image metrics between *FFT unwrap, FFT wide,* and *FFT nav* for the fourth echo (TE = 19 ms). This figure complements Figure 9 in the main text. Further details can be found in that figure caption.

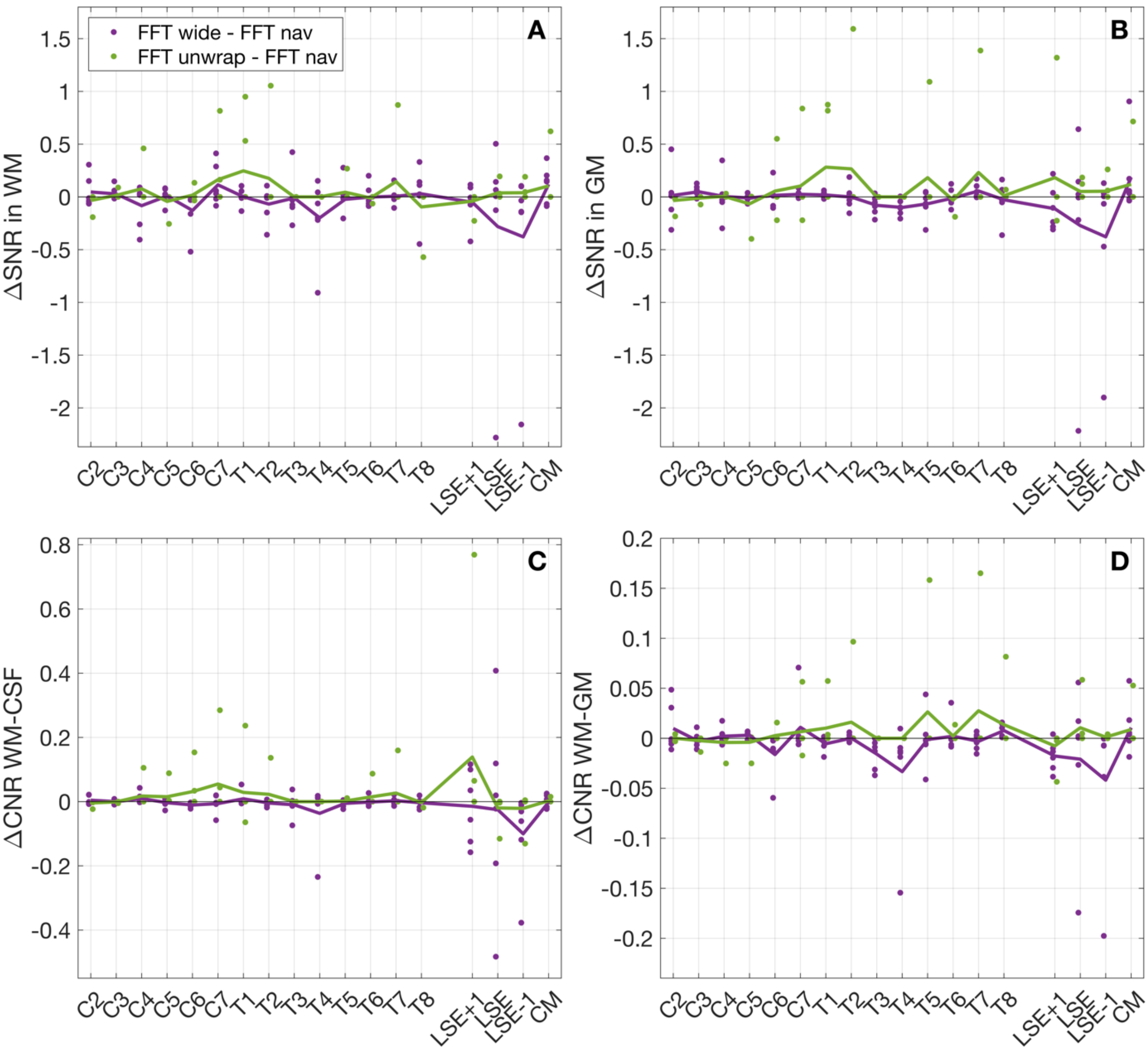


**Figure S5:** Comparison of image metrics between *FFT unwrap, FFT wide,* and *FFT nav* for the root-mean-square (RMS) images. This figure complements Figure S3. Further details can be found in that figure caption.

#### Discussion

##### The lungs as round spheres: field simulation

When an object with a given magnetic susceptibility distribution ($\chi(\vec{r})$) is located in an external magnetic field $\vec{B}_0 = B_0 \cdot \vec{z}$, the field is perturbed. The resulting field perturbation ($B$) can be calculated using a Fourier-based method to approximate the solution to the Maxwell equations[2,3]. This method assumes that the xy-plane components of the induced magnetization vector are negligible compared to the z component and that $|\chi| \ll 1$. According to this method each element in the magnetization distribution can be treated as an independent dipole $D(\vec{r})$, and the field perturbation in the z direction ($B_z$) can be computed as[2,3]

$$\frac{B_z(\vec{r})}{B_0} = \chi(\vec{r}) \otimes D_z(\vec{r}) \ ,$$

where $D_z(\vec{r})$ is the z component of the dipole kernel expressed as

$$D_z(\vec{r}) = \frac{1}{4\pi} \cdot \frac{3\cos^2(\theta) - 1}{|\vec{r}|^3} \ ,$$

where $\theta$ is the angle between $\vec{B}_0$ and $\vec{r}$. This equation is more conveniently solved in k-space where the convolution with the dipole kernel becomes a multiplication according to the convolution theorem.

This Fourier-based method was used to simulate the field perturbation due to the lungs in the surrounding region, when a subject is laying in the MR scanner. The lungs were modelled as two spherical objects with identical volume and positive magnetic susceptibility compared to the surroundings, as expected for air relative to soft tissues[3] (Figure S6 panel A). The field simulation was run two times using two different volumes for the spheres, mimicking inhalation and exhalation. The profile difference between inhalation and exhalation was then computed (Figure S6 panel C). The pattern of the simulated field fluctuations obtained with this model is consistent with the pattern of field fluctuations across vertebral levels measured in vivo[1,4] (Figure 4).

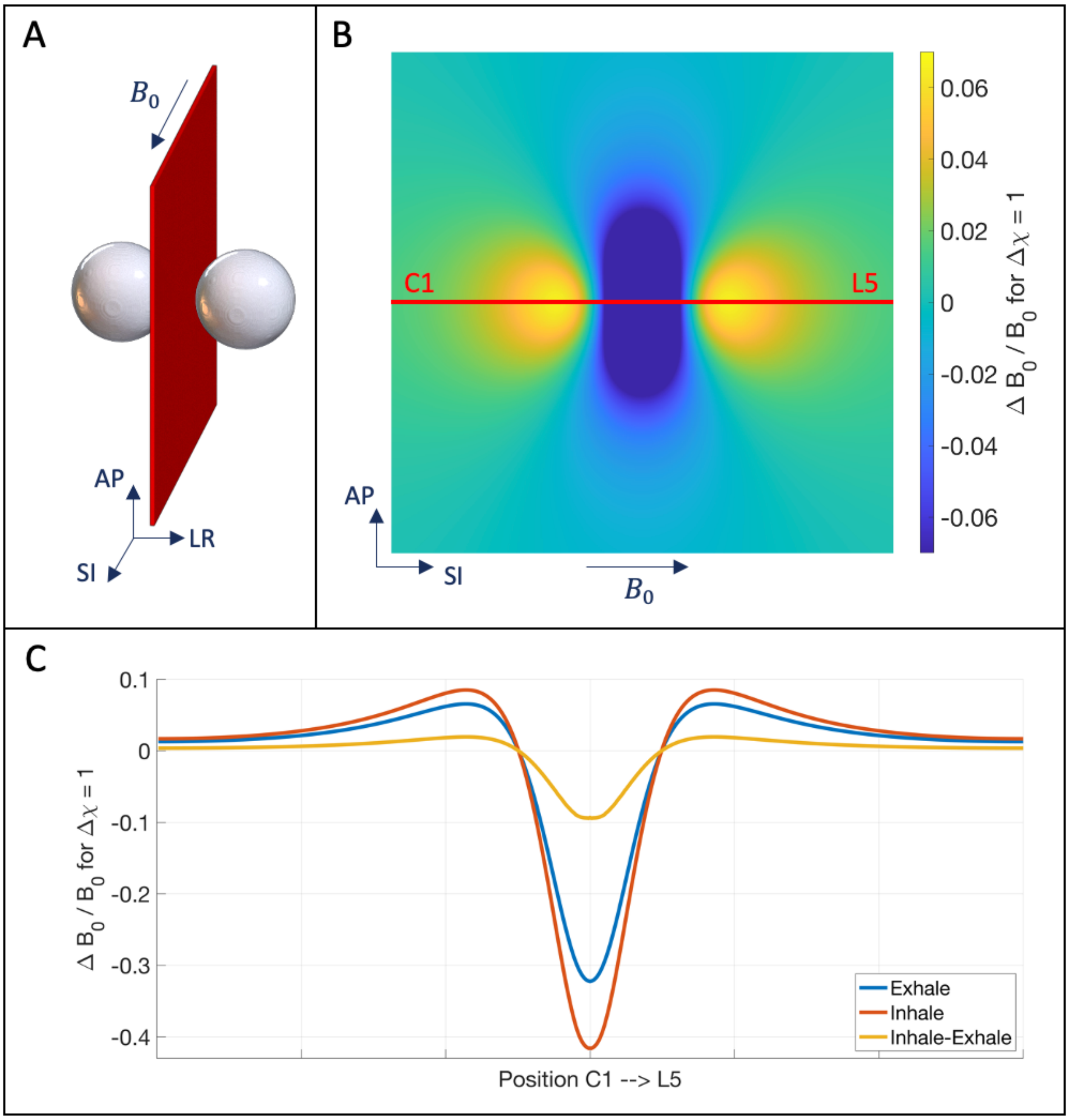


**Figure S6:** The lungs were modelled as two spheres in a magnetic field with positive susceptibility compared to the surroundings (panel A). Panel B shows the field perturbation due to the two spheres on the red plane in between the spheres. Panel C shows the field perturbation profile along the superior-inferior direction (corresponding to the red line on the map). The field simulation was run two times using two different volumes for the spheres, mimicking inhalation and exhalation. The profile difference between inhalation and exhalation was then computed and is shown in yellow.
